# Measurement Equivalence of the ASRS Across the Adult Lifespan: A Differential Item Functioning Analysis

**DOI:** 10.64898/2026.04.06.26350233

**Authors:** Noa Givon-Schaham, Nir Shalev

## Abstract

Adult ADHD is increasingly recognized across the lifespan, yet the psychometric equivalence of the Adult ADHD Self-Report Scale (ASRS) remains unverified for older populations. This study examined age-related Differential Item Functioning (DIF) in 600 adults (n = 100 per decade, ages 20– 80) who completed the 18-item ASRS. Using a bi-factor Graded Response Model, we extracted latent ADHD trait scores (ωH =.895) and assessed DIF via ordinal logistic regression with adaptive age modeling. Five of 18 items exhibited significant uniform DIF. At equivalent latent severity, older adults were less likely to endorse hyperactivity symptoms in Part A (fidgeting, feeling “driven by a motor”) but more likely to endorse specific symptoms in Part B (careless mistakes, misplacing items, interrupting). From ages 20 to 80, expected Part A scores decreased by 1.36 points (∼0.27 per decade), while Part B scores increased by 1.15 points (∼0.23 per decade). These findings indicate a phenotypic redistribution of ADHD symptoms as individuals age. Because the 6-item Part A screener serves as the primary clinical gatekeeper, its concentration of negative DIF suggests standard screening practice may systematically underestimate ADHD severity in older adults. We recommend using the full 18-item ASRS when screening older populations and suggest that developing age-adjusted norms would improve diagnostic accuracy.

## Introduction

Attention-Deficit/Hyperactivity Disorder (ADHD) is a prevalent neurodevelopmental syndrome characterized by difficulties in attention, hyperactivity, and impulsivity, often persisting from childhood into adulthood ^1^. ADHD remains largely underdiagnosed in older populations ^2^. Prevalence estimates decline from 5–7% in young adulthood to approximately 1–3% after age 50 ^3^, but this reduction is confounded by significant underdiagnosis, with a ten-fold discordance between symptom-based estimates and clinical diagnoses in older cohorts ^4^.

Several factors drive the underdiagnosis of ADHD in older age. Inattention and executive dysfunction are frequently misattributed to typical cognitive aging or psychiatric comorbidity ^5^, while overt hyperactivity tends to “mellow” into internal restlessness with age ^1^, yielding a predominantly inattentive phenotype difficult to distinguish from mild cognitive impairment ^6,7^. Diagnostic clarity is further hindered by cohort effects, specifically the lack of formal diagnostic recognition prior to 1980, which complicate the retrospective childhood validation required for diagnosis ^8^.

The Adult ADHD Self-Report Scale (ASRS ^9^) is the most widely used screening instrument in epidemiological and clinical settings. However, its original validation focused primarily on adults aged 18-44^1,9^, and no dedicated age-stratified psychometric analyses have established its performance in older adulthood. If ASRS items are interpreted differently across age groups, the instrument may introduce systematic measurement bias. The present study examined this possibility using Differential Item Functioning (DIF) analysis across ages 20–80, to assess whether the ASRS functions comparably across the adult lifespan and evaluate implications for ADHD screening in later life.

## Methods

### Ethics

The study was approved by the Ethics Committees at the University of Oxford (approval number R74132/RE004) and the University of Haifa (approval number 405/24).

### Participants and Procedure

The full procedural and demographic details are reported in Supplementary Materials (S1 and S2). Six hundred adults were recruited online in six age brackets of N = 100 each (20–29; 30–39; 40– 49; 50–59; 60–69; 70–80). Inclusion criteria required English proficiency, residence in the US or UK, normal or corrected-to-normal vision, no diagnosed psychiatric or neurological condition, and a minimum approval rating of 90% in previous experiments. The resulting cohort (281 men, 319 women, one non-binary) had a mean age of 50.15 years (SD = 17.3; range 20-80).

### Measure

ADHD symptom severity was assessed using the WHO Adult ADHD Self-Report Scale (ASRS-v1.1 ^9^; Kessler et al., 2005), an 18-item instrument mapping onto DSM-IV Criterion A symptoms. Part A comprises a 6-item screener; Part B a 12-item supplementary checklist. Items are rated from 0 (“Never”) to 4 (“Very Often”). As extended description of the ASRS is in Supplementary S3.

### Analysis

Extended details of the analysis procedure appear in S4, Supplementary Materials. All analyses were conducted in R. Response categories were collapsed from 5 to 4 points by merging “Often” and “Very Often” to address sparse endorsement among older adults. A Bi-factor Graded Response Model was estimated via Quasi-Monte Carlo EM, specifying a General ADHD factor loading on all 18 items orthogonal to Inattention (Items 1–4, 7–11) and Hyperactivity/Impulsivity (Items 5–6, 12–18) specific factors; individual latent trait scores (θ) were extracted using Expected A Posteriori estimation. DIF was assessed via Ordinal Logistic Regression, comparing baseline (θ only), Uniform DIF (θ + Age), and Non-Uniform DIF (θ + Age + θ×Age) models. Age was modelled with natural cubic splines (df = 2), with automatic fallback to a linear term when cell counts fell below N = 8. p-values were corrected using Benjamini-Hochberg. To quantify DIF impact, Expected Total Scores were computed separately for Part A and Part B, comparing participants aged 20 versus 80 at equivalent latent severity. Analysis code and data are available at OSF (https://osf.io/rwqtn/overview?view_only=f80fb62201644e5ca9ab45797b8e0b6c).

## Results

### Structural Validity and Reliability

Detailed results are in Table S1 (Supplementary Materials S5). The bi-factor model demonstrated acceptable global fit. The General ADHD factor showed high hierarchical reliability (ωH = .895) and accounted for most common variance (ECV = .802).

### Differential Item Functioning

Likelihood ratio tests revealed significant Uniform DIF in 5 of 18 items after Benjamini-Hochberg correction (all adjusted p < .002; full statistics in Table S2, Appendix 5). No items exhibited significant Non-Uniform DIF, indicating that item discrimination remained stable across age.

In Part A, two hyperactivity/impulsivity items showed negative age effects: Item 5 (“fidget or squirm”) and Item 6 (“overly active and compelled to do things”). Older adults were less likely to endorse these items than younger adults at equivalent latent ADHD severity. In Part B, three items showed positive age effects: Item 7 (“careless mistakes”), Item 10 (“difficulty finding things”), and Item 18 (“interrupt others when busy”). Here, older adults were more likely to endorse these symptoms at the same trait level. The expected item score curves for all five items are depicted in Figure 1.

**Figure 1:**
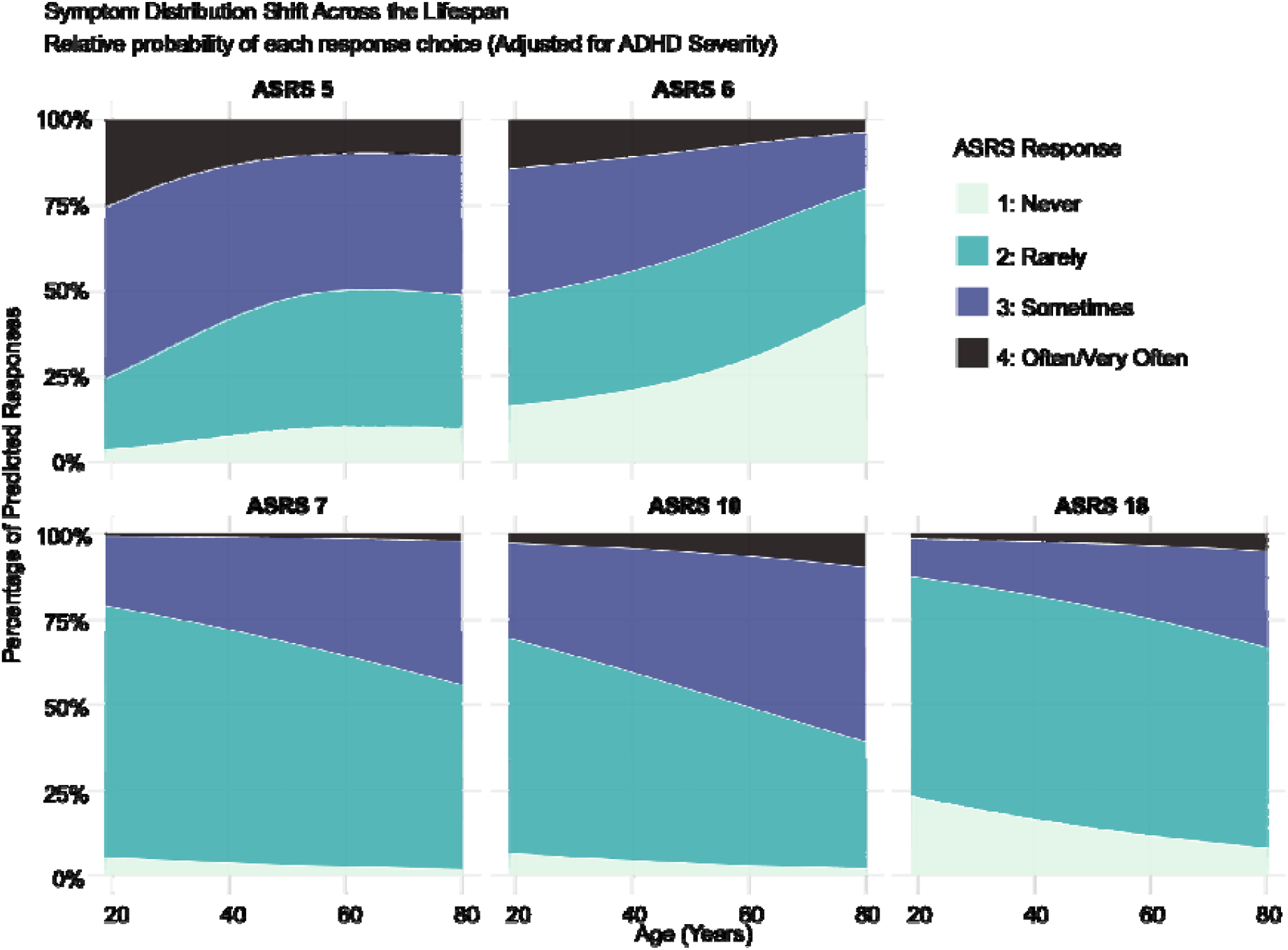
Expected item score curves for the five ASRS items exhibiting significant age-based Differential Item Functioning (DIF). The upper two rows depict symptoms from the Part A clinical screener (Items 5 and 6), which show a negative age effect; older adults are significantly less likely to endorse these symptoms than younger adults at the same level of latent ADHD severity. The bottom row display symptoms from the Part B checklist (Items 7, 10, and 18), which demonstrate a positive age effect: these symptoms are more readily endorsed by older adults. Each panel plots the expected raw score against the latent ADHD trait (θ), illustrating the “phenotypic redistribution” of symptoms across the lifespan.

To quantify the cumulative impact of DIF on clinical assessment, Expected Total Scores (ETS) were calculated for Part A and Part B at the mean latent severity (θ = 0), comparing participants aged 20 versus 80 (Figure 2). For Part A, older adults scored approximately 1.36 points lower than younger adults (∼0.27 points per decade decrease), suggesting the screener may systematically underestimate severity in older populations. For Part B, the pattern reversed: older adults scored approximately 1.15 points higher (∼0.23 points per decade increase), indicating these symptoms are more readily endorsed with age.

**Figure 2:**
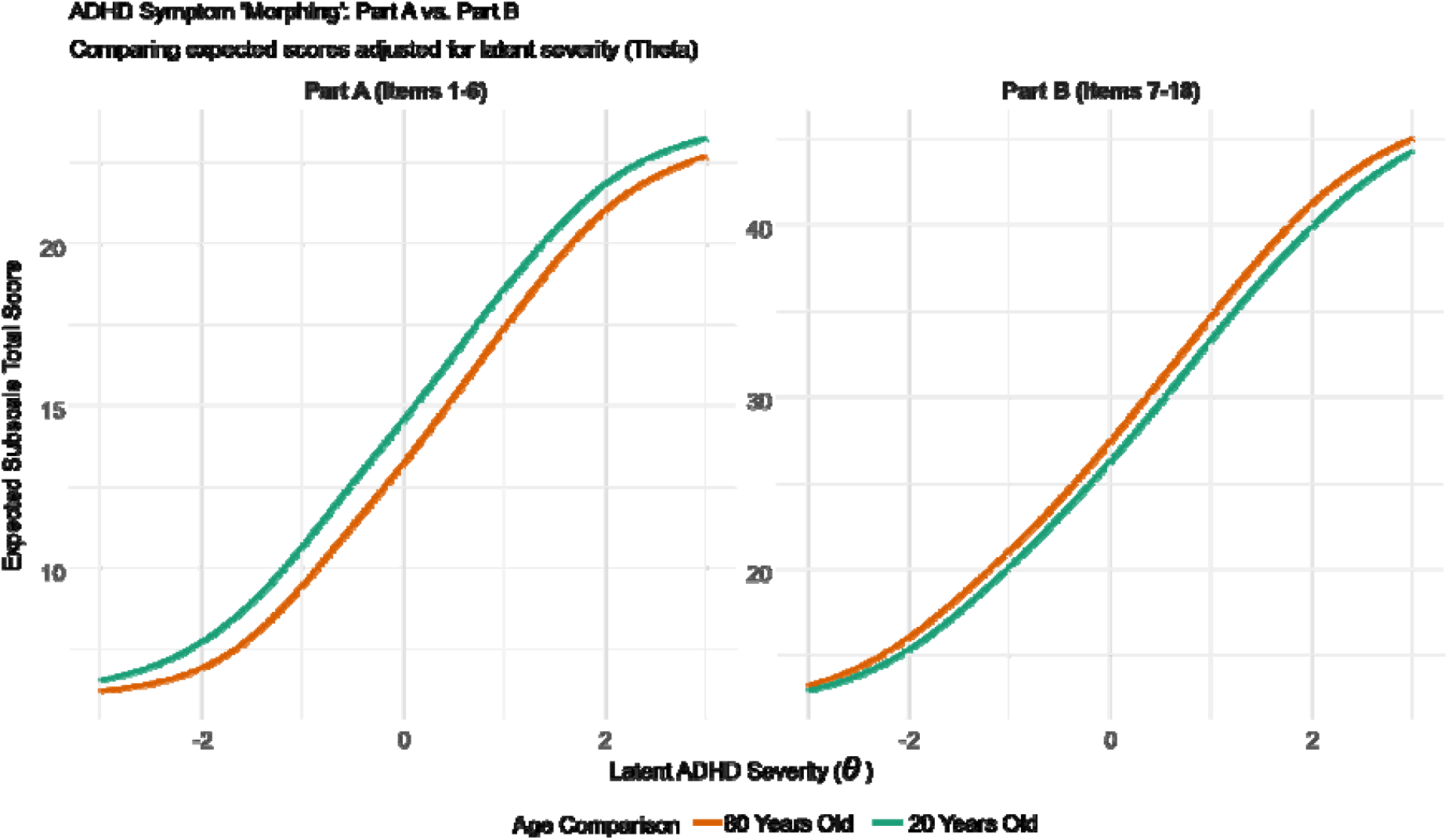
Expected Total Scores (ETS) for the ASRS Part A screener and Part B checklist as a function of age-related Differential Item Functioning. The left panel displays Part A (Items 1 to 6) and the right panel displays Part B (Items 7 to 18). The x-axis represents latent ADHD severity (θ), and the y-axis represents the expected subscale total score. Two curves are plotted in each panel: green for a 20-year-old and orange for an 80-year-old at equivalent levels of latent severity. For Part A, the green curve (age 20) sits above the orange curve (age 80), indicating that older adults receive lower screener scores than younger adults at the same underlying ADHD severity, with an estimated gap of approximately 1.36 points at θ = 0 (approximately 0.27 points per decade). For Part B, the pattern reverses: the orange curve (age 80) rises above the green curve (age 20), reflecting higher endorsement of inattentive and disorganised symptoms with age, with an estimated gap of approximately 1.15 points at θ = 0 (approximately 0.23 points per decade). These opposing trajectories illustrate a phenotypic redistribution of ADHD symptom expression across the lifespan, whereby the Part A screener increasingly underrepresents older adults’ true symptom burden while the full 18-item instrument provides a more balanced assessment.

### Divergence of Part A and Part B Trajectories

ASRS Part A showed a stronger negative correlation with age (r = −.26, p < .001) than Part B (r = −.12, p = .002). Steiger’s test for dependent overlapping correlations confirmed this difference was significant, t(597) = −4.85, p < .001 (Figure S1, supplementary materials S6).

## Discussion

The present findings demonstrate that the ASRS does not function equivalently across the adult lifespan. When younger and older adults share the same underlying level of ADHD severity, they do not receive the same scores. Specifically, older adults score lower on items capturing hyperactivity and impulsivity - fidgeting, feeling compelled to be active - while scoring higher on items reflecting inattentive and disorganized behaviors such as making careless mistakes, misplacing things, and interrupting others. In practical terms, two individuals with identical ADHD severity could receive meaningfully different scores depending on their age, not because their disorder differs, but because the instrument captures it differently.

These findings should be situated within the broader developmental trajectory of ADHD. Prevalence estimates decline substantially from young adulthood into later life (^3,4^). Our results suggest that at least part of this apparent change may reflect measurement issues: a shift in which and to what degree some of the symptoms are endorsed at different age groups. Critically, the age-related DIF we observed was not uniform across the instrument: it concentrated in specific items and operated in opposite directions across the two subscales. This asymmetry has direct clinical consequences.

The Part A screener, i.e., the 6-item subset most commonly used as a clinical gatekeeper, contained the two items with the strongest negative age bias. If clinicians or researchers rely solely on Part A, as is common in primary care and epidemiological screening, they risk systematically underestimating ADHD in older adults. The Part B results tell a different story: the age-related reduction in symptom endorsement is substantially weaker, and several items show increased endorsement with age. The full 18-item instrument thus provides a more balanced picture of ADHD severity across the lifespan. This screening bias may further obscure the clinical picture given the existing overlap between late-life ADHD and mild cognitive impairment ^6,7,10^.

Our results call for a reconsideration of how ADHD is screened in older adulthood. As a first and readily implementable step, clinicians and researchers should ensure that the full 18-item ASRS is used when assessing older adults, rather than relying on the 6-item Part A screener alone. More broadly, the field would benefit from developing age-adjusted norms or dedicated screening tools that account for the phenotypic redistribution of ADHD symptoms across the lifespan. Until such tools are available, awareness of these measurement biases is essential to avoid the continued under-identification of ADHD in older adulthood.

## Data Availability

All data produced are available online at (https://osf.io/rwqtn/overview?view_only=f80fb62201644e5ca9ab45797b8e0b6c).

## Acknowledgements

N.S. is supported by the Israeli Science Foundation Research Grant (1073/24).

## Supplementary materials

### S1. Extended Participants and Procedure

Participants were recruited via Prolific (www.prolific.co), a platform for recruiting participants for online experiments ^11^. Inclusion criteria involved proficiency in English, living in the US or the UK, having normal or corrected-to-normal vision, and no diagnosed psychiatric or neurological condition. Additionally, participants were required to have completed at least twenty previous studies on Prolific with a minimum approval rating of 90% or more (i.e., the percentage of studies on Prolific for which each participant’s data had been approved). This criterion was applied to increase the reliability of the data.

All participants used a personal computer to complete the study (the experiment did not support phones or tablets). Participants provided informed consent and were compensated at a rate of £9 per hour, in line with Prolific’s recommendation. Participants logged in to Prolific and used a link that directed them to a survey running on the Pavlovia platform for online studies (https://pavlovia.org/). They signed a consent form and then completed a demographic questionnaire and the Adult ADHD Self-Report Scale (ASRS-v1.1 ^9^). The questionnaires were generated using PsychoPy ^12^.

Data were pooled from two independent experimental studies. Wave 1 (N = 300) was conducted as part of a study on spatial attention ^13^, while Wave 2 (N = 300) was part of a separate battery of tasks investigating visual working memory. These additional cognitive procedures also served as a data quality check: after completing the first round of recruitment in each wave, participants who did not complete the entire study or performed on the cognitive task more than 2 SD below the sample mean (and always above chance level) were replaced.

To ensure data comparability, the demographic questionnaire and ASRS were administered at the start of both sessions, prior to any experimental manipulations. Procedural consistency was maintained across both waves, and participants from Wave 1 were excluded from participating in Wave 2 to ensure a fully independent combined sample of N = 600.

### S2. Demographic Details

The sample included 600 participants in six age brackets of N = 100 each: 20–29, 30–39, 40–49, 50– 59, 60–69, and 70–80. The resulting cohort comprised 281 men, 319 women, and one non-binary participant, with a mean age of 50.15 years (SD = 17.3; range 20–80).

In Wave 1 (N = 300; mean age = 50.51; SD = 17.33; range 20–80), only information on age, sex, and years of education was collected (mean years of education = 16.13; SD = 6.6). Wave 2 (N = 300; mean age = 49.8; SD = 17.27; range 20–80) included more detailed background and demographic information. The ethnic composition of the Wave 2 sample was as follows: 8.33% identified as African American, 2.33% as Asian, 2% as Hispanic or Latino, 0.33% as North African, 83.33% as White/Caucasian, and 2.66% did not disclose their identity. Regarding educational attainment, 40% held a Bachelor’s degree, 23% had a high school education, 11% held a Master’s degree, 8.66% had a professional diploma, 12.33% had completed secondary school, and 4.33% did not disclose.

### S3. ASRS Questionnaire

ADHD symptom severity was assessed using the World Health Organization (WHO) Adult ADHD Self-Report Scale (ASRS-v1.1) Symptom Checklist. Developed as part of the WHO World Mental Health Survey Initiative, this 18-item instrument maps directly onto the Criterion A symptoms of ADHD as defined in the DSM-IV ^9^. The scale is structured into two components: Part A, a 6-item screener consisting of the most predictive symptoms; and Part B, a 12-item checklist providing additional clinical elaboration. Participants rated the frequency of each symptom over the past six months on a 5-point Likert scale ranging from 0 (“Never”) to 4 (“Very Often”). The instrument has demonstrated high internal consistency and, notably, high sensitivity in primary care settings ^14^.

Although the diagnostic framework has since transitioned to the DSM-5, the symptom content remains comparable between the two editions. The primary modification in the DSM-5 was a reduction in the clinical threshold for adults, requiring five endorsed symptoms rather than the six required under DSM-IV. Because the ASRS-v1.1 measures continuous symptom frequency rather than categorical diagnostic status, it remains a valid instrument for assessing severity under both classifications ^15^. The ASRS-v1.1, which captures the full range of Criterion A symptoms, remains a widely accepted measure of continuous ADHD severity.

### S4. Extended Analysis Details

#### Data Preparation and Category Management

All analyses were conducted using R statistical software (Version 4.x). The analysis pipeline consisted of three stages: (1) data preparation and sparsity management, (2) latent trait estimation using a high-stability bi-factor model, and (3) Differential Item Functioning (DIF) analysis using an adaptive ordinal regression framework. The analysis code and the experimental data are fully available at OSF https://osf.io/rwqtn/overview?view_only=f80fb62201644e5ca9ab45797b8e0b6c.

Prior to analysis, response patterns were inspected for sparsity, particularly within the upper age quantiles. Preliminary analysis revealed that the highest response category (“Very Often”) was endorsed with extremely low frequency by older adults on specific items (e.g., ASRS-5, ASRS-18), leading to structural zeros that destabilised ordinal likelihood estimation. To address this and ensure robust convergence across the lifespan, the response scale was collapsed from a 5-point to a 4-point scale by merging the two highest categories (“Often” and “Very Often”) into a single “High Endorsement” category. This collapsing strategy was applied uniformly across all items and participants to preserve measurement consistency.

#### Latent Trait Estimation

To account for the multidimensional structure of the ASRS-18, we utilised a Bi-factor Graded Response Model (GRM). The bi-factor structure specified a single General ADHD Factor (G) loading on all 18 items, orthogonal to two specific factors: Inattention (Items 1–4, 7–11) and Hyperactivity/Impulsivity (Items 5–6, 12–18). Given the complexity of estimating high-dimensional bi-factor models with ordinal data, we employed Quasi-Monte Carlo Expectation-Maximisation (QMC-EM) estimation via the mirt package ^16^.

QMC-EM was selected over standard EM algorithms to ensure global convergence and stability of the standard errors, with the number of estimation cycles increased to 5,000 to prevent premature termination. The individual latent trait scores (θ) for the General Factor were extracted using the Expected A Posteriori (EAP) method. This general factor served as the matching variable for subsequent DIF analyses, ensuring that comparisons between age groups were adjusted for true underlying ADHD severity.

#### Differential Item Functioning (DIF) Analysis

DIF was assessed using a Ordinal Logistic Regression implemented in the ordinal package in R ^17^. This method models the cumulative probability of endorsing a response category as a function of the latent trait (θ) and the covariate of interest (Age). For each item, three nested models were compared:

Model 0 (Baseline): Item response predicted solely by Latent ADHD Severity (θ). Model 1 (Uniform DIF): Item response predicted by θ + Age. Model 2 (Non-Uniform DIF): Item response predicted by θ + Age + (θ × Age).

Adaptive Model Specification: To capture potential non-linear developmental shifts (e.g., symptoms peaking in middle age), Age was initially modelled using Natural Cubic Splines (df = 2). However, to address methodological concerns regarding sparsity in the oldest age brackets (70+), an adaptive algorithm was implemented. If the minimum cell count for an item-age quadrant fell below N = 8, the spline term was automatically replaced with a linear Age term. This adaptive approach prevented overfitting and false-positive “Caution” flags caused by model instability in sparse tails. Hypothesis Testing: Uniform DIF (a systematic shift in severity thresholds across age) was tested by comparing Model 1 vs. Model 0 via a Likelihood Ratio Test (LRT). Non-Uniform DIF (a difference in item discrimination across age) was tested by comparing Model 2 vs. Model 1. To control the False Discovery Rate (FDR) due to multiple comparisons (18 items × 2 tests), p-values were adjusted using the Benjamini-Hochberg (BH) procedure.

#### Assessment of DIF Impact and Visualisation

To evaluate the practical magnitude of DIF, we conducted two impact analyses. First, we generated predicted probability curves for all response categories as a function of age, holding latent severity constant (θ). These plots visualise how the definition of “symptom severity” shifts developmentally (e.g., whether older adults shift from “Often” to “Sometimes” for the same underlying trait level). Second, we calculated the Expected Total Score (ETS) for the ASRS Part A (Screener) and Part B (Symptom Checklist) separately. By comparing the ETS of a 20-year-old versus an 80-year-old at identical levels of latent severity (θ), we quantified the “Score Gap”, i.e., the number of points “lost” or “gained” solely due to age bias.

### S5. Extended DIF Results

Individual item-level DIF statistics are reported below. Items reaching significance after Benjamini-Hochberg correction are presented in bold.

**Table S1.** Standardised Bi-factor Loadings for the ASRS-18.

| Item | General (G) | Inatt (S1) | Hyp/Imp (S2) | Communality (h<sup>2</sup>) |
| --- | --- | --- | --- | --- |
| ASRS 1 | 0.834 | 0.332 | — | 0.805 |
| ASRS 2 | 0.855 | 0.412 | — | 0.901 |
| ASRS 3 | 0.697 | 0.176 | — | 0.517 |
| ASRS 4 | 0.805 | 0.234 | — | 0.703 |
| ASRS 5 | 0.683 | — | −0.052 | 0.470 |
| ASRS 6 | 0.511 | — | 0.298 | 0.350 |
| ASRS 7 | 0.668 | −0.187 | — | 0.481 |
| ASRS 8 | 0.639 | −0.403 | — | 0.571 |
| ASRS 9 | 0.660 | −0.244 | — | 0.496 |
| ASRS 10 | 0.591 | −0.095 | — | 0.358 |
| ASRS 11 | 0.588 | −0.217 | — | 0.393 |
| ASRS 12 | 0.634 | — | 0.268 | 0.474 |
| ASRS 13 | 0.706 | — | 0.182 | 0.531 |
| ASRS 14 | 0.623 | — | 0.158 | 0.413 |
| ASRS 15 | 0.566 | — | 0.525 | 0.596 |
| ASRS 16 | 0.534 | — | 0.548 | 0.585 |
| ASRS 17 | 0.645 | — | 0.515 | 0.680 |
| ASRS 18 | 0.622 | — | 0.476 | 0.613 |
| **ωH** | **0.895** |  |  |  |
| **ECV** | **0.802** |  |  |  |

*Note.* G = General ADHD factor; S1 = Inattention specific factor; S2 = Hyperactivity/Impulsivity specific factor. Dashes indicate items not loading on that specific factor. ωH = Omega Hierarchical; ECV = Explained Common Variance.

**Table S2.** Differential Item Functioning Statistics for All 18 ASRS Items.

| Item | Model Type | Uniform $\chi^2$ (df) | p value | Non-Unif $\chi^2$ (df) | p value | Effect |
| --- | --- | --- | --- | --- | --- | --- |
| 1 | Linear | 0.02 (1) | .924 | 1.34 (1) | .800 | N.S. |
| 2 | Linear | 0.33 (1) | .730 | 0.03 (1) | .919 | N.S. |
| 3 | Linear | 0.01 (1) | .924 | 0.55 (1) | .800 | N.S. |
| 4 | Spline (df=2) | 5.41 (2) | .172 | 1.10 (2) | .800 | N.S. |
| **5** | **Spline (df=2)** | **15.83 (2)** | **.002** | **0.48 (2)** | **.919** | **Decreasing** |
| **6** | **Spline (df=2)** | **28.98 (2)** | **<.001** | **1.40 (2)** | **.800** | **Decreasing** |
| **7** | **Linear** | **11.81 (1)** | **.002** | **2.72 (1)** | **.445** | **Increasing** |
| 8 | Spline (df=2) | 0.53 (2) | .862 | 1.47 (2) | .800 | N.S. |
| 9 | Linear | 0.93 (1) | .547 | 0.73 (1) | .800 | N.S. |
| **10** | **Linear** | **18.21 (1)** | **<.001** | **0.86 (1)** | **.800** | **Increasing** |
| 11 | Linear | 1.15 (1) | .511 | 0.05 (1) | .919 | N.S. |
| 12 | Linear | 0.79 (1) | .560 | 7.63 (1) | .104 | N.S. |
| 13 | Spline (df=2) | 4.98 (2) | .186 | 2.59 (2) | .800 | N.S. |
| 14 | Spline (df=2) | 0.63 (2) | .862 | 0.28 (2) | .919 | N.S. |
| 15 | Spline (df=2) | 1.58 (2) | .630 | 6.79 (2) | .302 | N.S. |
| 16 | Spline (df=2) | 4.01 (2) | .269 | 0.00 (2) | .998 | N.S. |
| 17 | Linear | 4.70 (1) | .090 | 3.80 (1) | .308 | N.S. |
| **18** | **Linear** | **17.29 (1)** | **<.001** | **0.37 (1)** | **.800** | **Increasing** |

*Note.* All p-values are adjusted using the Benjamini-Hochberg procedure. Significant items (adjusted $p < .05$ ) are presented in bold. “Decreasing” = negative age effect (lower endorsement by older adults); “Increasing” = positive age effect (higher endorsement by older adults). Model type indicates whether age was modelled using natural cubic splines (df = 2) or a linear term, determined by the adaptive algorithm (see S4.3).

### S6. Supplementary Figure

**Figure S1**. Divergence of Part A and Part B age–score correlations. ASRS Part A (6-item screener) showed a significantly stronger negative correlation with age (r = −.26, p < .001) than Part B (12-item checklist; r = −.12, p = .002). The difference was confirmed by Steiger’s test for dependent overlapping correlations, t(597) = −4.85, p < .001.

**Figure S1:**
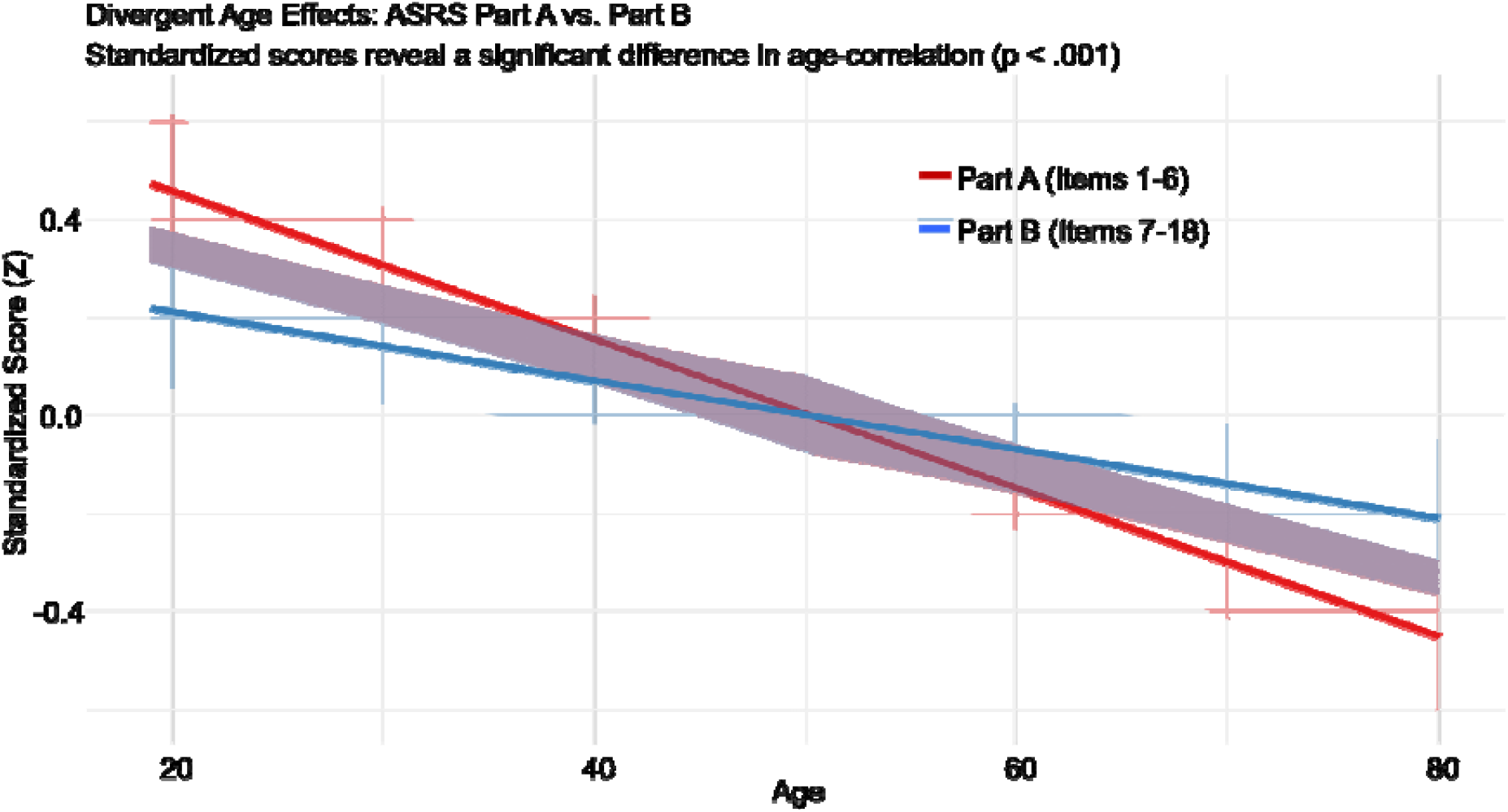
Divergent age effects on ASRS Part A and Part B standardised scores. The x-axis represents participant age (ranging from 20 to 80 years), and the y-axis represents standardised total scores (Z). Regression lines with shaded 95% confidence bands are shown separately for Part A (Items 1 to 6; red) and Part B (Items 7 to 18; blue). Both subscales show a negative linear trend with age, but the slope is markedly steeper for Part A. Part A exhibited a stronger negative correlation with age (r = -.26, p < .001) than Part B (r = -.12, p = .002), and the difference between these dependent correlations was confirmed by Steiger’s test for dependent overlapping correlations, t(597) = -4.85, p < .001. This divergence is consistent with the Differential Item Functioning results reported in the main text: the concentration of negatively biased hyperactivity/impulsivity items in Part A drives a steeper age-related decline in screener scores, while the presence of positively biased inattention items in Part B partially offsets the overall decline, yielding a shallower slope for the supplementary checklist.

